# Exploring the causal role of the human gut microbiome in colorectal cancer: Application of Mendelian randomization

**DOI:** 10.1101/2022.10.14.22281077

**Authors:** Charlie Hatcher, George Richenberg, Samuel Waterson, Long H. Nguyen, Amit D. Joshi, Robert Carreras-Torres, Victor Moreno, Andrew T. Chan, Marc Gunter, Yi Lin, Conghui Qu, Mingyang Song, Graham Casey, Jane C Figueiredo, Stephen B Gruber, Jochen Hampe, Heather Hampel, Mark A Jenkins, Temitope O Keku, Ulrike Peters, Catherine M Tangen, Anna H Wu, David A Hughes, Malte C Rühlemann, Jeroen Raes, Nicholas J. Timpson, Kaitlin H. Wade

## Abstract

**Aim:** The role of the human gut microbiome in colorectal cancer (CRC) is unclear as most studies on the topic are unable to discern correlation from causation. We apply two-sample Mendelian randomization (MR) to estimate the causal relationship between the gut microbiome and CRC.

**Materials and methods:** We used summary-level data from independent genome-wide association studies to estimate the causal effect of 14 microbial traits (n=3,890 individuals) on overall CRC (55,168 cases, 65,160 controls) and site-specific CRC risk, conducting several sensitivity analyses to understand the nature of results.

**Results:** Initial MR analysis suggested that a higher abundance of *Bifidobacterium* and presence of an unclassified group of bacteria within the *Bacteroidales* order in the gut increased overall and site-specific CRC risk. However, sensitivity analyses suggested that instruments used to estimate relationships were likely complex and involved in many potential horizontal pleiotropic pathways, demonstrating that caution is needed when interpreting MR analyses with gut microbiome exposures. In assessing reverse causality, we did not find strong evidence that CRC causally affected these microbial traits.

**Conclusions:** Whilst our study initially identified potential causal roles for two microbial traits in CRC, importantly, further exploration of these relationships highlighted that these were unlikely to reflect causality.

## Introduction

Colorectal cancer (CRC) is the third most common cause of cancer death worldwide, with over 16,000 deaths due to CRC every year in the UK alone(1,2). Whilst research suggests that over half of CRC cases are likely explained by preventable causes(1,3), CRC remains an ever-increasing burden, particularly among young people. Therefore, it is important to identify novel modifiable risk factors to help reduce the global incidence of, and deaths from, CRC.

There is increasing evidence that the human gut microbiome – the naturally occurring complex community of microorganisms housed within the gastrointestinal tract – plays a role in human health, including influencing the risk and prognosis of CRC. As the gut microbiome has a substantial impact on host metabolism(4,5), inflammation(6,7), and host immune response to both commensal and pathogenic microbes(7,8), there are many plausible biological mechanisms by which the gut microbiome could influence cancer development(4,9,10). However, findings within this context have been inconsistent and unclear, making it difficult to draw conclusive evidence. For example, whilst there have been some compelling results from *in vivo* and *in vitro* studies showing a reduced incidence of CRC through modification of single or a small consortia of microbial constituents (e.g., with pre- or pro-biotics)(11), few of these findings have been successfully translated between model organisms and into humans harbouring more complex communities.

Despite this emerging evidence, studies generating this body of work have been unconvincing in their ability to offer causal evidence given their observational nature and lack of temporality. For example, evidence has suggested an overall lower diversity in microbiota in cases of CRC compared to controls in addition to lower levels of *Bifidobacterium* and *Roseburia* spp. and higher levels of *Fusobacterium* and *Porphyromonas* bacteria (12–14). However, these existing epidemiological studies are usually cross-sectional or case-control designs and have a limited ability to discern correlation from causation. In addition, they often have small sample sizes, limiting power to detect associations. Furthermore, human randomized controlled trials (RCTs) have been unable to conclusively show strong evidence for a reduction in the incidence of CRC using treatments designed to alter the gut microbiome(15). Furthermore, discrepancies in the literature are likely due to the challenges in multi-omic technologies (e.g., sequencing and metagenomics), sensitive experimental models and important limitations of conventional epidemiological studies such as confounding, reverse causation and bias. Evidence that has not translated between model organisms impedes opportunity for harnessing the gut microbiome for improving population health(16).

In the absence of large-scale RCTs, Mendelian randomization (MR) is a method that offers improved causal inference by utilizing human germline genetic variation (usually, single nucleotide polymorphisms [SNPs]) as instruments for a clinically relevant trait (here, the gut microbiome) in a manner that is analogous to RCTs to mitigate some of the biases present within conventional epidemiological studies(17–20). As germline genetic variation is randomly inherited and fixed at conception, results of MR analyses should be largely independent from confounding and traditional reverse causation (i.e., the outcome leading to variation in the exposure). There is an increasing literature utilising MR to assess the causal implications of variation in the gut microbiome (using host genetic variation associated with gut microbial composition) on various health outcomes. However, there is an unmet requirement for careful examination and interpretation of these estimates given the potential complexity of host (i.e., human) genetic effects on the gut microbiome, despite their ability to yield apparently causal estimates. In this study, we utilised and explored the properties of two-sample MR analyses based on host microbiome genetic effects to interrogate causal relationships between variation in the human gut microbiome (specifically, 14 microbial traits representing either relative abundances or the likelihood of presence versus absence of certain bacteria) on overall and site-specific CRC risk.

## Results

### Effects of gut microbial traits on CRC

We utilised GWAS summary statistics to perform two-sample MR to estimate the causal effect of the gut microbiome on overall CRC and four CRC sites (distal, proximal, colon, and rectal) (see Methods, **Supplementary Table 1** and **Figure 1**). Genetic variants associated with 14 microbial traits were obtained from a microbiome GWAS (mGWAS) meta-analyses(21) conducted in 3,890 individuals. Specifically, 13 SNPs exceeding a genome-wide meta-analysis threshold (p< 5×10^−08^) were selected as genetic instruments for 13 microbial traits (with each microbial trait instrumented by a single SNP), in addition to one SNP associated with bacteria of the *Bifidobacterium* genus that has previously consistently been reported in the literature. All F-statistics were greater than 31 (**Supplementary Table 2**). The variance explained by each associated SNP in each microbial trait ranged between 0.8% (the genus, *Veillonella*) and 1.6% (the unclassified group of bacteria in the *Porphyromonadaceae* family).

**Figure 1.**
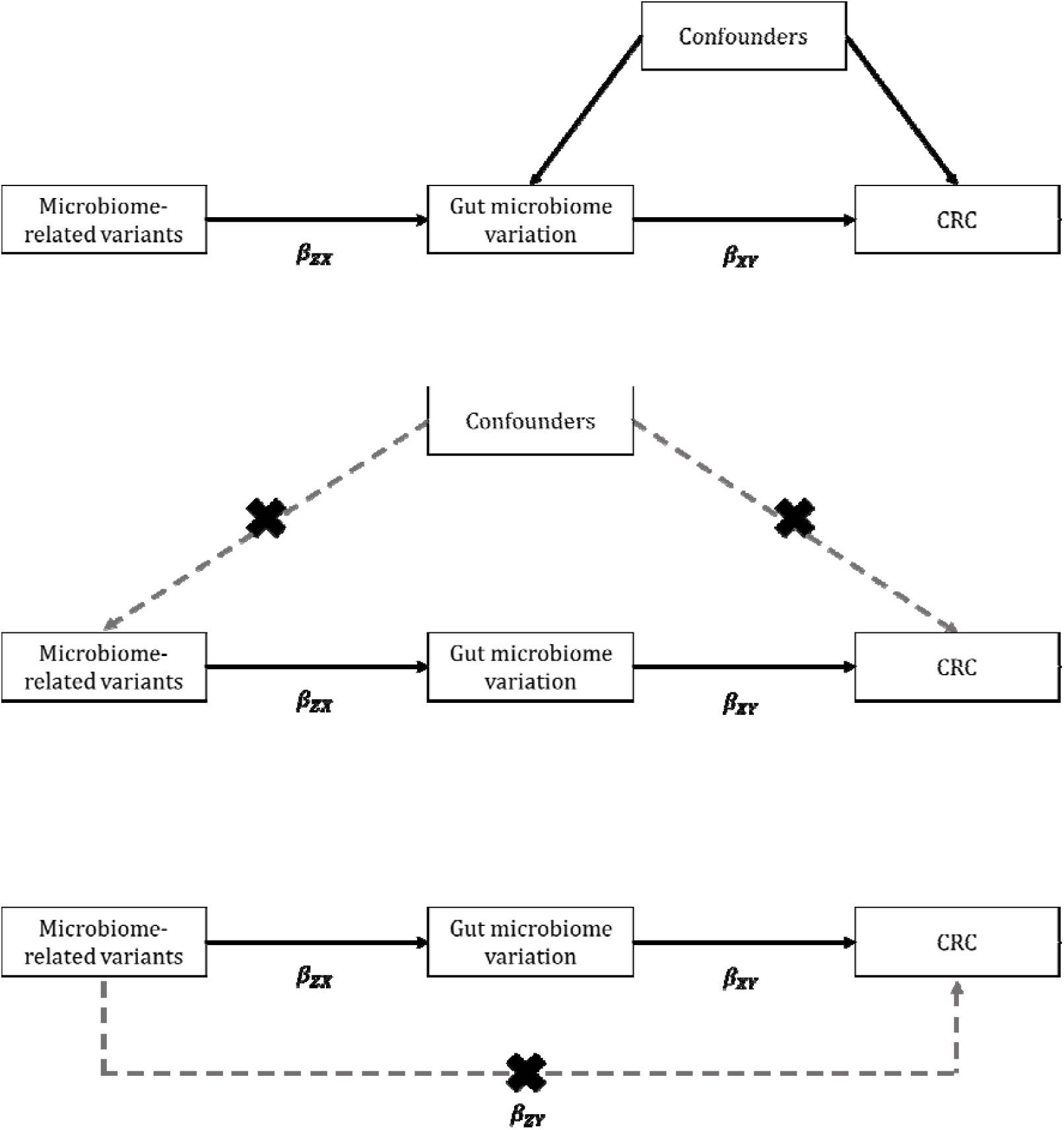
Mendelian randomization framework applied to assess the causal effect of the human gut microbiome on CRC risk. Abbreviations: CRC = colorectal cancer; MR = Mendelian randomization; SNP = single nucleotide polymorphism. MR relies on three key assumptions: (top panel) the SNPs are associated with the exposure (here, the gut microbiome); (middle panel) there are no common causes of the SNPs and the outcome (here, CRC), meaning any confounders driven by population substructure, dynastic effects or assortative mating (these may not and are unlikely to be the same confounders of the association between the exposure and outcome); and (bottom panel) the SNPs are not independently associated with the outcome (here, CRC) other than pathways through the exposure (here, the gut microbiome). Given these key assumptions, microbiome-related variants can be used to assess the causal effect of the human gut microbiome on CRC, overcoming limitations of observational epidemiological studies. In two-sample MR analyses, the causal effect of the exposure on outcome (β_XY_) is generated by a ratio of the SNP-outcome (β_ZY_) and the SNP-exposure (β_ZX_) effect estimates derived from two independent samples.

In main MR analyses, there was evidence for a causal role of a higher abundance of bacteria within the *Bifidobacterium* genus (*G. Bifidobacterium*) increasing the risk of CRC by approximately 41% (odds ratio [OR] per standard deviation [SD] higher relative abundance: 1.41; 95% CI: 1.20, 1.65, P=1.83×10^−05^; **Figure 2**). The presence (versus absence) of an unclassified group of bacteria within the *Bacteroidales* order (*G. unclassified, O. Bacteroidales*) was found to increase the risk of CRC (OR per approximate doubling of the genetic liability to presence versus absence: 1.08; 95% CI: 1.02, 1.16; P=0.01). Similar relationships were observed between these two microbial traits and the four CRC sites studied (**Table 1**). Specifically, a higher abundance of *Bifidobacterium* increased the risk of all four CRC sites: distal colon cancer (OR: 1.61; 95% CI: 1.26, 2.07; P=1.40×10^−04^), proximal colon cancer (OR: 1.31; 95% CI: 1.03, 1.67; P=0.03), colon cancer (OR: 1.40; 95% CI: 1.16, 1.69; P=3.75×10^−04^) and rectal cancer (OR: 1.61; 95% CI: 1.26, 2.05; P=1.16×10^−04^). Results for the unclassified group of bacteria within the *Bacteroidales* order also showed consistent directions of effect for all CRC sites: distal colon cancer (OR: 1.04; 95%: 0.93, 1.15; P=0.51), proximal colon cancer (OR: 1.12; 95% CI: 1.02, 1.24; P=0.02), colon cancer (OR: 1.09; 95% CI: 1.01, 1.18; P=0.03) and rectal cancer (OR: 1.11; 95% CI: 1.00, 1.23; P=0.05). However, confidence intervals (CIs) for distal colon cancer were wide, likely due to the smaller sample size. MR effect estimates for the remaining 12 microbial traits on overall and site-specific CRC risk were much smaller in size and had 95% CIs that crossed the null (**Table 1**).

**Table 1.**
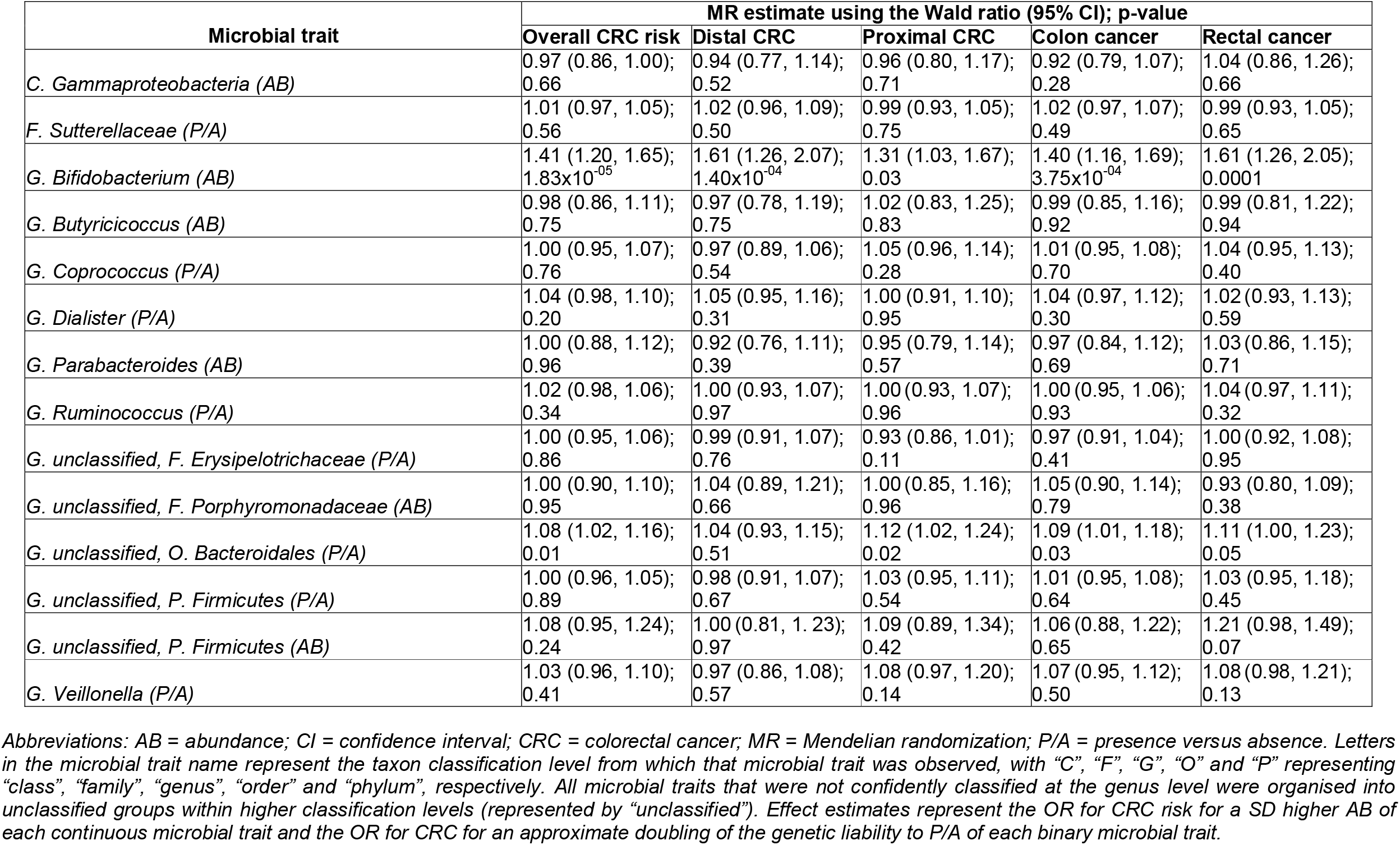
Two-sample Mendelian randomization estimates of the effect of each microbial trait on overall and site-specific CRC risk.

**Figure 2.**
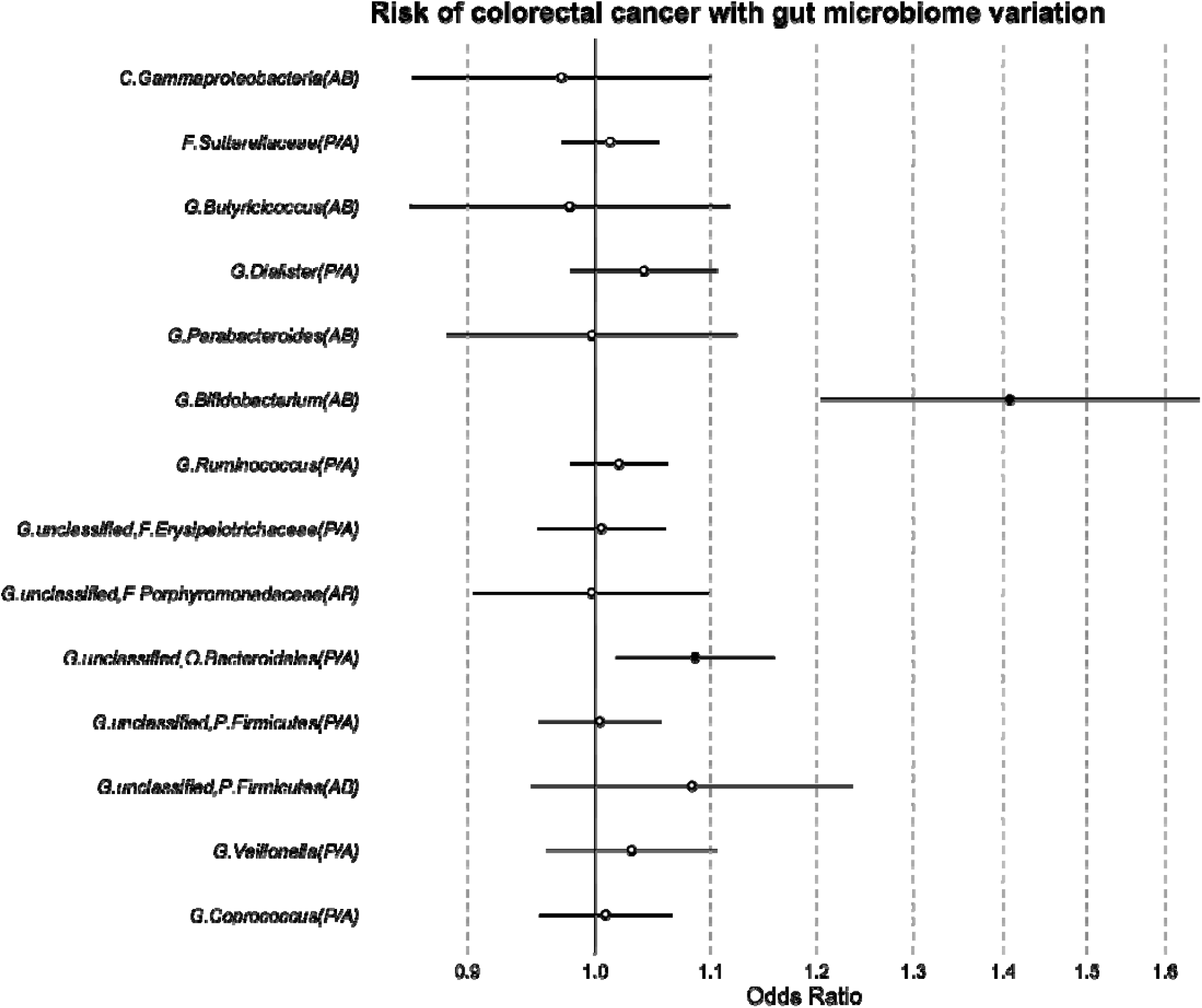
MR estimates of the effect of each microbial trait on overall CRC risk. Abbreviations: AB = abundance; CI, confidence interval; CRC = colorectal cancer; MR = Mendelian randomization; OR = odds ratio; P/A = presence versus absence; SD = standard deviation. MR estimates represent the OR for CRC risk and 95% CI per SD unit change for continuous microbial traits (labelled as “AB”) or per approximate doubling of the genetic liability to presence (vs. absence) of each binary microbial trait (labelled as “P/A”).

### Colocalisation

We performed genetic colocalisation to determine whether the single shared genetic variant being used as an instrument for each microbial trait was associated with both variation in that microbial trait and CRC. Specifically, we performed colocalisation on the two microbial traits which showed strong evidence of a causal effect in the MR analyses (*G. Bifidobacterium* and *G. unclassified, O. Bacteroidales*) with overall and site-specific CRC (distal CRC was not considered in the colocalisation analyses with *G. unclassified, O. Bacteroidales* since evidence for a causal effect was weaker in the main MR analyses). Genome-wide data for microbial traits were only available from the Flemish Gut Flora Project (FGFP) GWAS limiting analyses to 2223 participants. Additionally, unlike in the mGWAS meta-analysis, neither lead SNP (rs4988235 and rs116135844) reached genome-wide significance in the FGFP dataset. Therefore, focusing on the difference between the tested hypotheses (see Methods), colocalisation results provided evidence that neither *G. Bifidobacterium* nor *G. unclassified, O. Bacteroidales* were likely to share a causal variant with overall or site-specific CRC (**Table 2**).

**Table 2.**
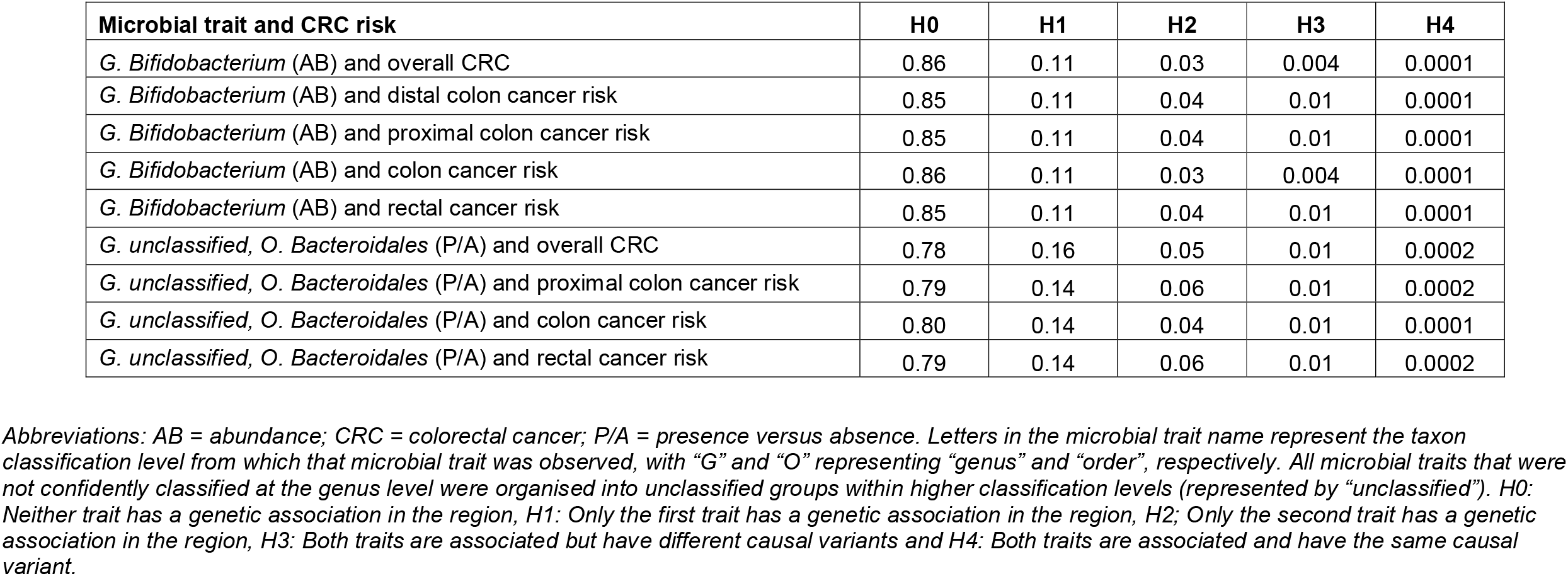
Posterior probabilities relating to associations between microbial traits and colorectal cancer subtypes.

| **Microbial trait and CRC risk** | **H0** | **H1** | **H2** | **H3** | **H4** |
| --- | --- | --- | --- | --- | --- |
| *G. Bifidobacterium* (AB) and overall CRC | 0.86 | 0.11 | 0.03 | 0.004 | 0.0001 |
| *G. Bifidobacterium* (AB) and distal colon cancer risk | 0.85 | 0.11 | 0.04 | 0.01 | 0.0001 |
| *G. Bifidobacterium* (AB) and proximal colon cancer risk | 0.85 | 0.11 | 0.04 | 0.01 | 0.0001 |
| *G. Bifidobacterium* (AB) and colon cancer risk | 0.86 | 0.11 | 0.03 | 0.004 | 0.0001 |
| *G. Bifidobacterium* (AB) and rectal cancer risk | 0.85 | 0.11 | 0.04 | 0.01 | 0.0001 |
| *G. unclassified, O. Bacteroidales* (P/A) and overall CRC | 0.78 | 0.16 | 0.05 | 0.01 | 0.0002 |
| *G. unclassified, O. Bacteroidales* (P/A) and proximal colon cancer risk | 0.79 | 0.14 | 0.06 | 0.01 | 0.0002 |
| *G. unclassified, O. Bacteroidales* (P/A) and colon cancer risk | 0.80 | 0.14 | 0.04 | 0.01 | 0.0001 |
| *G. unclassified, O. Bacteroidales* (P/A) and rectal cancer risk | 0.79 | 0.14 | 0.06 | 0.01 | 0.0002 |

Abbreviations: AB = abundance; CRC = colorectal cancer; P/A = presence versus absence. Letters in the microbial trait name represent the taxon classification level from which that microbial trait was observed, with “G” and “O” representing “genus” and “order”, respectively. All microbial traits that were not confidently classified at the genus level were organised into unclassified groups within higher classification levels (represented by “unclassified”). H0: Neither trait has a genetic association in the region, H1: Only the first trait has a genetic association in the region, H2: Only the second trait has a genetic association in the region, H3: Both traits are associated but have different causal variants and H4: Both traits are associated and have the same causal variant.

Regional association plots confirmed findings from genetic colocalisation for *G. unclassified, O. Bacteroidales* (**Figure 3**; **Supplementary Figure 1**). However, despite little evidence for genetic colocalisation for *G. Bifidobacterium* with overall and site-specific CRC, regional association plots suggest the genomic region surrounding the rs4988235 SNP is important in both abundance of *G*.*Bifidobacterium* and CRC risk (**Figure 3**; **Supplementary Figure 2**).

**Figure 3.**
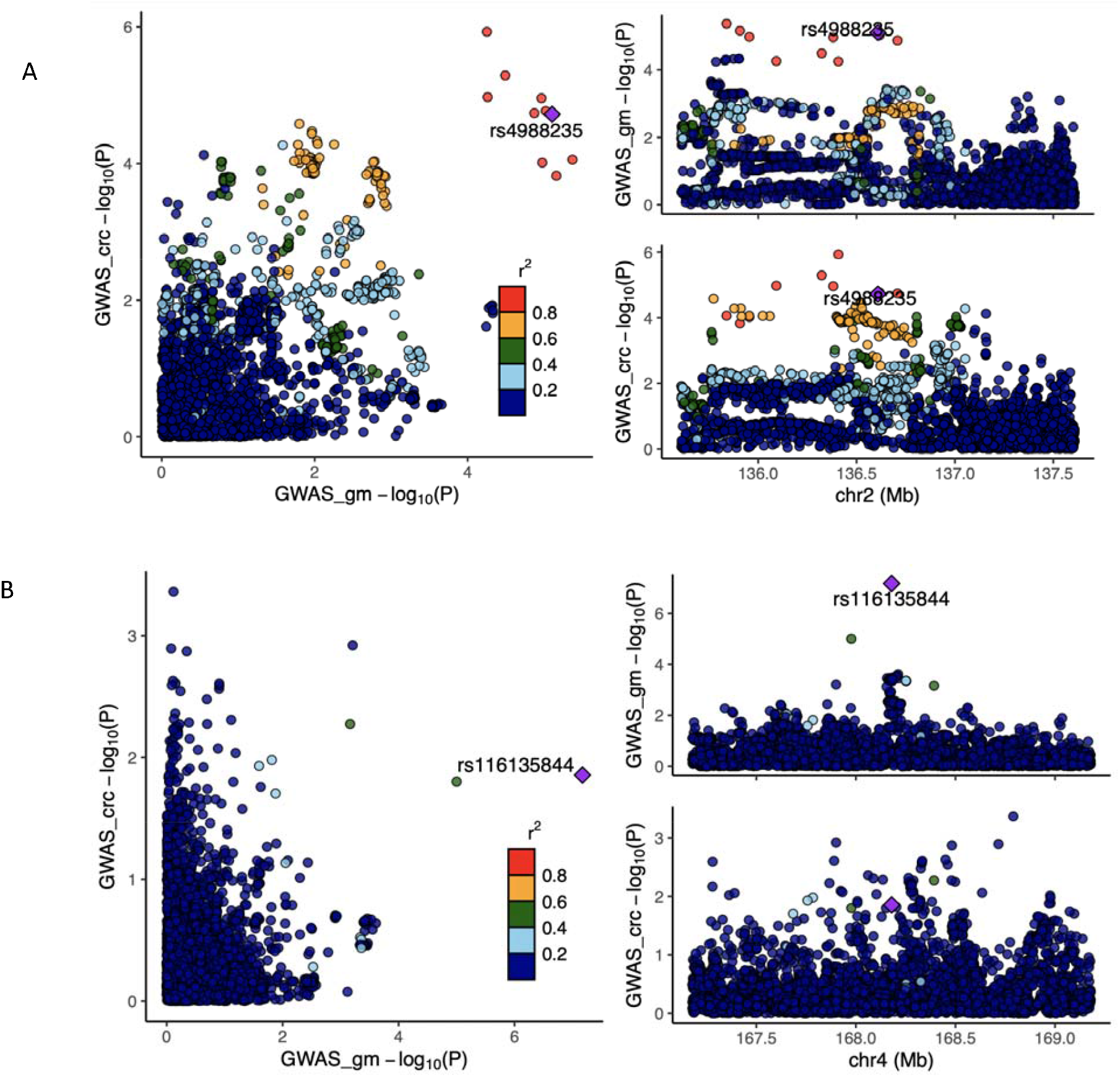
Colocalisation results for bacteria within the (A) *Bifidobacterium* AB microbial trait and (B) P/A of unclassified genera within the *Bacteroidales* order with overall CRC. Abbreviations: AB = abundance; CRC = colorectal cancer; FGFP = Flemish Gut Flora Project; GECCO = Genetics and Epidemiology of Colorectal Cancer Consortium; GM = gut microbiome; GWAS = genome-wide association study; P/A = presence versus absence. Regional association plots, generated from LocusCompareR, showing the -log10(P-value) where each lead SNP is represented by a purple diamond (panel A: rs4988235 associated with the Bifidobacterium AB microbial trait and panel B: rs116135844 associated with the unclassified Bacteroidales P/A microbial trait) in relation to overall CRC. These plots were created using the FGFP and GECCO full summary-level data for microbial traits and CRC, respectively.

### Manual exploration of pleiotropy

We searched summary-level GWAS data collated by PhenoScanner(22) and the IEU Open GWAS(23) to identify whether SNPs used to instrument *G. Bifidobacterium* and *G. unclassified, O. Bacteroidales* had previously been reported to be associated with CRC or any other trait that could be an independent cause of CRC. Searching GWAS summary statistics collated by PhenoScanner found evidence for potential horizontal pleiotropy between the SNP used as an instrument for *G. Bifidobacterium* (rs4988235) and CRC risk. A total of 51 associations with complex traits and diseases were observed at a defined multiple testing p-value threshold (P<1×10^−04^, see Methods) including positive associations with numerous anthropometric measures (i.e., fat mass, body mass index and waist circumference). Additionally, the rs4988235 SNP has also previously been associated with 103 expression quantitative traits (eQTs), 6 metabolites, 59 methylation quantitative traits (mQTs) and one protein (**Supplementary Tables 3-7**). In the IEU Open GWAS, over 230 traits were identified as being associated with rs4988235 at the same p-value threshold (**Supplementary Table 8**). Given the number of associations between this SNP and both complex and molecular traits, there are likely many plausible pathways between this SNP and CRC, which could be independent of the *G. Bifidobacterium*.

In contrast, we found very little evidence for horizontal pleiotropy between the SNP used as an instrument for the *G. unclassified, O. Bacteroidales* (rs116135844) and CRC risk, with only three associations observed at the pre-defined p-value threshold (P<1×10^−04^; **Supplementary Tables 9-12**). These traits included disorders of the patella, which is less likely to be biologically linked to CRC and therefore unlikely to reflect horizontal pleiotropy between the instrument and outcome. The other traits implicated in relation to rs116135844 included gene expression of the *UBE2J2* gene and an uncharacterized genetic probe (2490351). The *UBE2J2* gene encodes a member of the E2 ubiquitin-conjugating enzyme family that modifies abnormal proteins or short-lived proteins targeted for degradation. The GWAS Catalog showed various associations with *UBE2J2*, including many cardiometabolic measures (e.g., body mass index, systolic blood pressure and coronary heart disease), autoimmune diseases (e.g., inflammatory bowel diseases and ulcerative colitis), CRC and others, suggesting *UBE2J2* gene expression could have a downstream impact on CRC. Further to this, no associations of this SNP with any phenotypes in the IEU Open GWAS met our p-value threshold (**Supplementary Table 13**).

### Two-sample MR using a lenient p-value threshold for selection of genetic instruments

When using a more lenient p-value threshold (P<1×10^−05^) and restricting to SNPs with directionally consistent effect estimates across the three studies comprising the gut microbiome GWAS (**Supplementary Table 14**), nine microbial traits had additional associated SNPs that could be used as theoretical instruments for further MR analyses. Focusing on those microbial traits for which there was evidence of a causal effect on CRC, *G. Bifidobacterium* had five associated SNPs and *G. unclassified, O. Bacteroidales* had three associated SNPs in total. Inverse variance weighted (IVW)-derived estimates for the causal effect of these microbial traits on overall CRC were consistent in direction to those obtained in the main analyses; however, the magnitude of all estimates was attenuated, and CIs spanned the null (**Figure 4, Supplementary Table 15**). Specifically, a higher abundance of *G. Bifidobacterium* and presence of *G. unclassified, O. Bacteroidales* increased the risk of overall CRC by 16% (OR: 1.16; 95% CI; 0.99, 1.33; P=0.10) and 5% (OR: 1.05; 95% CI; 0.95, 1.14; P=0.34), respectively. Results for other microbial traits were consistent with the main analyses and provided little evidence for a causal effect on CRC risk.

**Figure 4.**
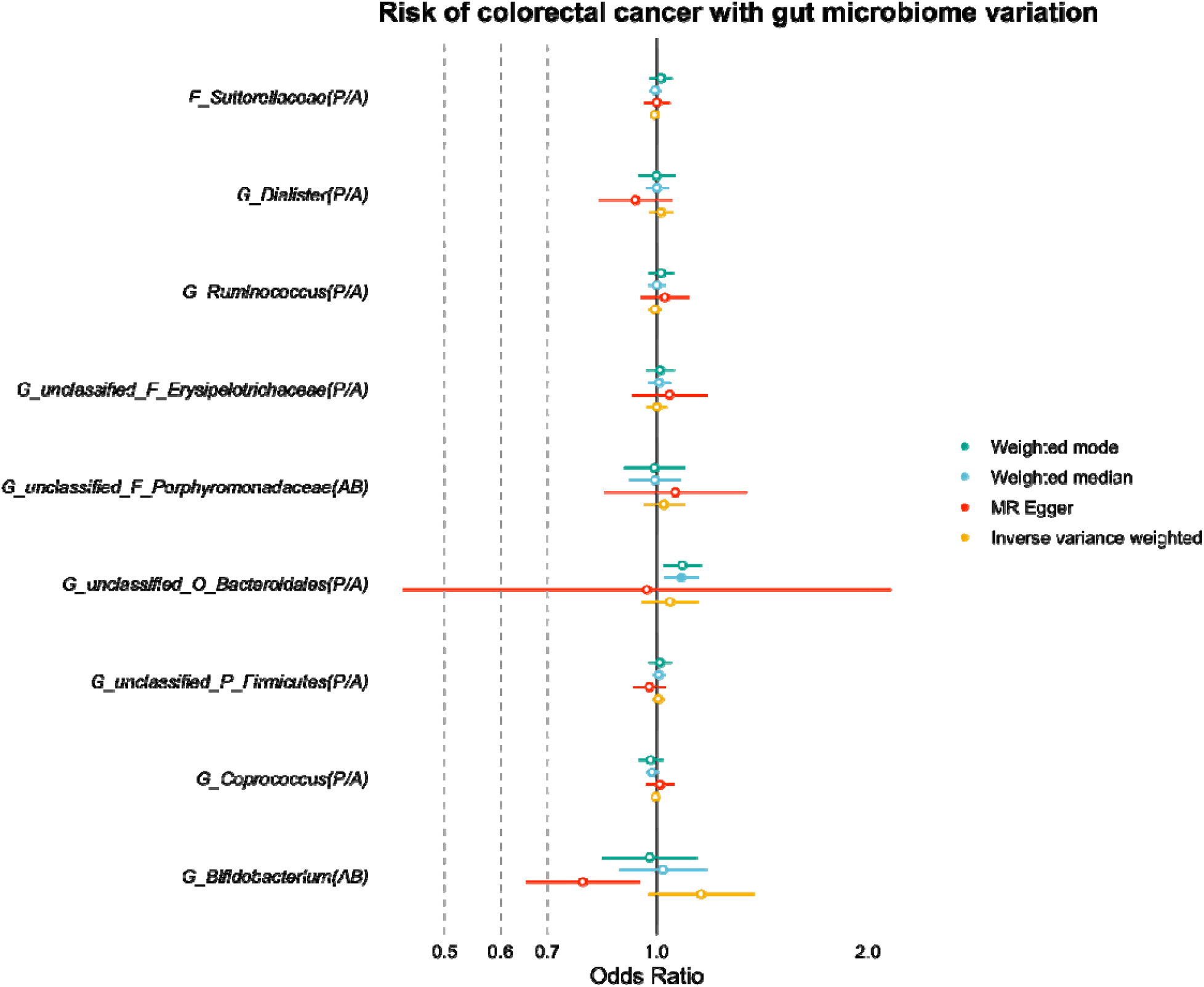
MR estimates of the effect of each microbial trait on overall CRC risk. Abbreviations: AB = abundance; CI = confidence interval; CRC = colorectal cancer; MR = Mendelian randomization; OR = odds ratio; P/A = presence versus absence; SD = standard deviation. MR estimates represent the OR for CRC risk and 95% CI per SD unit change for AB microbial traits or per approximate doubling of the genetic liability to P/A of each binary microbial trait. Results for the inverse variance weighted, weighted median, weighted mode and MR-Egger methods) are presented when using the multi-SNP instrument for each microbial trait using a lenient p-value threshold (P<1×10^−5^) and compared to the Wald ratio estimates obtained from the main analysis.

Estimates of the causal effect of *G. Bifidobacterium* on overall CRC risk using the pleiotropy-robust methods were inconsistent with those derived using the Wald ratio and IVW methods, where the weighted median estimate was positive but both the MR-Egger- and weighted mode-derived estimates were in the inverse direction (MR-Egger OR: 0.79; 95% CI: 0.60, 0.97; P=0.09 and weighted mode OR: 0.98; 95% CI: 0.82, 1.14; P=0.80). For *G. unclassified, O. Bacteroidales*, all estimates using pleiotropy-robust methods were consistent in direction suggesting that presence of this group of bacteria increased the risk of overall CRC; however, CIs were predictably wide for the MR-Egger effect estimate (**Figure 4, Supplementary Table 15**).

For site-specific analyses, IVW-derived effect estimates for the effect of *G. Bifidobacterium* were directionally consistent with the main analyses using the Wald ratio, though some estimates had wide CIs. The estimates for the causal effect of *G. Bifidobacterium* on distal, proximal and colon cancer were also consistent; however, results using the MR-Egger method were inverse for all sites of CRC with wide CIs. The estimates for the effect of *G. unclassified, O. Bacteroidales* on distal, proximal and colon cancer risk using the IVW and pleiotropy-robust methods were all directionally consistent (though with wide CIs) with main analyses, suggesting presence of this bacteria also increased the risk of site-specific CRC. However, estimates for the effect of both microbial traits on rectal cancer were inconsistent across all models, with the IVW and weighted median-based estimates being positive (and thus consistent with the main analyses) but either/both of the MR-Egger and weighted-mode-based estimates were inverse. Overall, these results may indicate that the originally observed causal effects in main analyses may be driven by horizontal pleiotropy (and downstream effects), which is also supported by our manual exploration of possible horizontal pleiotropic pathways.

### Reverse MR

To examine whether the causal effects identified between microbial traits and CRC risk were either bi-directional in nature or driven by reverse causation of the microbial trait-associated SNPs (i.e., the “microbial trait-related” SNP actually having a direct or indirect effect on CRC, which, in turn, actually impacts microbial trait variation), we performed reverse two-sample MR analyses. These analyses aimed to assess whether overall and site-specific CRC risk had a causal effect on *G. Bifidobacterium* and *G. unclassified, O. Bacteroidales* microbial traits. Results gave little evidence that overall CRC or CRC sites had a strong causal effect on either microbial trait (**Supplementary Table 16**).

## Discussion

In this study, we performed analyses to explore the causal role played by the gut microbiome and CRC. Whilst our initial MR analyses provided evidence that bacteria within the *Bifidobacterium* genera and an unclassified group of bacteria in the *Bacteroidales* order may increase the risk of overall and site-specific CRC, our sensitivity analyses highlighted that these relationships, and the tools used to assess them, are complex and, certainly in the case of *Bifidobacterium*, are unlikely to reflect causality. Our study therefore highlights the need for performing sensitivity analyses exploring the nature of the instruments and derived estimates when attempting to establish the causal role of the gut microbiome in health and disease.

Firstly, there was strong evidence for pleiotropy in the relationship between *G. Bifidobacterium* and both overall and site-specific CRC risk, where there were comfortably over 200 other traits that had previously been associated with the rs4988235 SNP that could be independently associated with CRC. Furthermore, when using multiple SNPs at a more lenient p-value threshold, the direction of effect was inconsistent across pleiotropy-robust methods. In reverse MR analyses, there was also no evidence to suggest a reverse effect in that a greater genetic liability to CRC was unlikely to drive variation in *Bifidobacterium* abundance. Therefore, the initially observed finding suggesting that a higher abundance of bacteria with the *Bifidobacterium* genus was likely not to reflect causality and, instead, reflect horizontal pleiotropy or a complex (potentially gene-environment) interaction between the genomic region surrounding the rs4988235 SNP (i.e., the *LCT/MCM6* gene) with both *Bifidobacterium* abundance and CRC risk, as indicated by our colocalisation analyses.

Conversely, our main results suggesting that the presence of an unclassified group of bacteria within the *Bacteroidales* order increased risk of CRC seemed not to be driven entirely by horizontal pleiotropy, as there were only a small number of plausible traits associated with the rs116135844 SNP that could be independently associated with CRC. In addition, results when using a more lenient p-value threshold were consistent with those from our main analyses. There was also no evidence to suggest a reverse effect in that a genetic liability to CRC was unlikely to change the greater genetic liability to presence of this unclassified group of *Bacteroidales* bacteria. Indeed, these findings support previous observational evidence suggesting that bacteria within the *Bacteroidales* order are more present in CRC cases compared to controls(24–26). There are also many existing studies showing a relationship between various *Bacteroides* species including *Bacteroides fragilis* and other species including *B. dorei, B. vulgatus* and *B. massilensis* with CRC(27). Whilst we were unable to classify the specific bacteria within this unclassified *Bacteroidales* microbial trait, these bacteria could plausibly contribute to the effect observed in the current analyses. Therefore, research using more granular measures of the gut microbiome (e.g., those capturing species- and strain-level bacteria or those able to characterize functionality such as metagenomics) will help provide clarity on these results specifically. Nevertheless, these results provide further evidence that the previously reported association between *Bacteroidales* genera and CRC may reflect causality.

However, it is worth noting that, whilst limited by power, our colocalisation analyses provided little evidence to suggest that either of these microbial traits (i.e., *G. Bifidobacterium* and *G. unclassified, O. Bacteroidales*) shared causal variants with CRC, which is a necessary (but not a sufficient) criteria for causality. Therefore, further analyses with a much larger quantity of microbiome-related SNPs, larger GWASs from which these SNPs are discovered and importantly replicated, plus a better understanding of the relationships between host genetic variation and the gut microbiome are required to provide conclusive evidence in this context.

Unlike the majority of the current studies applying MR to understand the causal implications of the gut microbiome and various health outcomes, the main strength of this study is that the microbiome-related SNPs used as instruments for the microbial traits are some of the first persistent signals across multiple cohorts(21). Whilst acknowledging that instrumentation of the gut microbiome is complex, we used genetic variants that were robustly related to each microbial trait (i.e., those reaching a genome-wide p-value threshold in the mGWAS). Compared to existing studies of this kind (which tend to use a lenient p-value threshold of 1×10^−05^ as a main analysis), this reduces the likelihood of including invalid SNPs within MR instruments. We did, however, opt to use more lenient p-value threshold for selection of genetic instruments (1×10^−05^) as a sensitivity analysis but importantly restricted SNPs to those which had a consistent direction of effect across multiple studies under the assumption that, in a larger GWAS, these SNPs would have reached a traditional genome-wide p-value threshold. Together, this provides more confidence that these are biologically relevant SNPs or at least those that were consistently associated with the microbial traits in this analysis. Additionally, GECCO is the largest GWAS of CRC to date, meaning that we were very well powered to detect a modest association in this current MR analysis (i.e., assuming an alpha level of 0.05 and an R^2^ of 0.01, we were 89% powered to detect an odds ratio of 1.2 in CRC). Generally, our study has shown the importance of performing several sensitivity analyses to assess the robustness of MR findings.

Despite this, there are several limitations to this work that centre around the core assumptions of the MR framework. Firstly, there must be no pathway between genetic instruments and the outcome (here, CRC) independent of the microbial exposure. Given that very little is known about the biology of the host genetic effects on the gut microbiome, there is a greater likelihood that the observed GWAS “microbiome-related” signals are reflective of either host-driven effects upstream of the gut microbiome (i.e., reverse causation in an MR context) or independent associations between these host genetic variants and, in this case, CRC (i.e., horizontal pleiotropy). This will remain a substantial limitation within the field without further large-scale GWASs of the gut microbiome and inter-disciplinary analyses clarifying the mechanistic pathways explaining the relationships between host genetic variation and gut microbiome variation.

Secondly, there are many microbiota of lower taxonomic units within the group of unclassified genera in the *Bacteroidales* order, emphasising a need to classify the exact species or strain of bacteria that could be driving this relationship and understand the mechanisms by which this occurs. Finally, even if these early results do suggest causality (with the case of *G. unclassified, O. Bacteroidales*), we do not currently know whether attempts to alter microbiota to reduce CRC risk would have other, unforeseen effects on other aspects of health. This is particularly important for the inclusion of the gut microbiome in the development of targeted therapeutics or preventative strategies for CRC.

Our study initially provided evidence that individuals who have a higher abundance of bacteria within the *Bifidobacterium* genus and presence of an unclassified group of bacteria within the *Bacteroidales* order within the gut may be at an increased risk of CRC and its sites. However, through exploration of invalidation of MR assumptions, we performed numerous sensitivity analyses which showed that these relationships are complex and may not reflect direct or causal relationships. Therefore, caution is required when interpreting these results and MR estimates alone are insufficient to determine causality in these contexts. Inter-disciplinary collaboration and triangulation across multiple, complementary study designs are required to understand the causal mechanisms that link both host genetic variation with the gut microbiome and the relationships between the gut microbiome and human health.

## Materials and methods

### Study design

Two-sample MR was used to examine the causal relationship between features of the human gut microbiome and both overall and site-specific CRC risk. In brief, SNP-exposure and SNP-outcome associations were obtained from independent non-overlapping GWASs to generate causal effect estimates of 14 microbial traits on overall and site-specific CRC risk. This study has been conducted in line with the Strengthening the Reporting of Observational Studies in Epidemiology MR (STROBE-MR) reporting guidelines for MR studies(28).

### Gut microbiome GWAS data and instrument selection

Genetic variants associated with microbial traits were obtained from one of the largest microbiome GWAS (mGWAS) meta-analyses of bacterial abundance, presence (versus absence), alpha- and beta-diversity and enterotype conducted within the FGFP (n=2,223) and two independent validation cohorts (the Food-Chain Plus study [FoCus, n=950] and the PopGen study [n=717])(21). Full details of the data sampling, preparation and analyses have been described previously(21,29,30). Briefly, DNA was extracted from frozen fecal samples provided by participants and hypervariable regions of the 16S rRNA gene were amplified and sequenced (the V4 hypervariable region for FGFP and the V1-V2 hypervariable regions for both FoCus and PopGen). Resulting sequences were analysed per sample using the DADA2 pipeline to provide taxonomical classifications of all bacteria in each sample(31). Bacterial classifications that were not confidently assigned at the genus level were organised into arbitrary unclassified groups within higher classification levels. Informed consent and appropriate ethical approval were obtained for each study.

In the FGFP cohort, which was used the discovery cohort in the GWAS analysis, the DADA2 pipeline yielded 499 taxonomical units across five levels of microbiota phylogeny from phylum to genus. Four metrics on alpha-diversity (within-individual diversity) and beta-diversity (inter-individual differences) were estimated using data for all 288 genus-level taxa. An enterotype phenotype, describing the different phylum-level community compositions, was also estimated.

After performing taxa-level quality control (QC), 92 taxa were available for the mGWAS (details previously described(21)). Given the ecological count nature of the 16S rRNA data, many microbial traits had zero-inflated distributions, which is problematic for linear modelling. Of the 92 taxa, 62 contained substantial zero-inflation; therefore, these taxa were modelled using a two-step hurdle binary analysis, which includes a binary presence (versus absence) analysis (denoted throughout as “P/A”) and a zero-truncated rank normal transformed abundance analysis (denoted throughout as “AB”). All other taxa were relatively normally distributed; therefore, were treated as AB phenotypes and rank normal transformed accordingly. Thus, a total of 159 microbial traits (i.e., 62 P/A, 92 AB, 3 alpha-diversity, 1 beta-diversity and 1 enterotype phenotypes) were analysed in the mGWAS conducted in the FGFP cohort.

Individuals from the FGFP cohort were genotyped on the Human Core Exome v1.0 and the Human Core Exome v1.1. Imputation was conducted using the 1000 Genomes data (phase 3) as the reference panel. After variant-level and individual-level QC followed by imputation, 7,711,310 variants and 2259 individuals remained, 2223 of whom also had data on the gut microbiome and all covariates used in the mGWAS analysis. All individuals were of European ancestry.

All mGWAS analyses were adjusted for extraction type and year, aliquot year, person performing the aliquot, library preparation plate, the first 10 genetic principal components, sex and age. Assuming an additive genetic model and accounting for genotype uncertainty, all AB and alpha-diversity microbial traits were regressed on covariates and residuals were regressed on genotype probability data in univariate linear models, all P/A microbial traits were analysed using a multivariate logistic model, all beta-diversity microbial traits were analysed using a multivariate model and the enterotype microbial trait was analysed using a multinomial logistic regression for categorial traits. All SNPs that reached an inclusive association test p-value threshold of 1×10^−05^ in the FGFP data set (n=23,735) were taken forward into a targeted meta-analysis including the independent FoCus and PopGen studies. Three genera were not present in these German cohorts; therefore, after excluding both P/A and AB microbial traits in these three instances (i.e., six microbial traits in total), the meta-analysis was limited to 153 microbial traits and 23,067 SNPs.

The FoCus and PopGen cohorts were genotyped using the Illumina Omni Express + Exome array and the Affymetrix Genome-Wide Human SNP Array 6.0, respectively. Imputation was conducted using the Sanger Imputation Service with the Human Reference Consortium (HRC) version 1.1 as the reference panel. The mGWAS meta-analyses were performed using the inverse-variance fixed effects method, with SNPs considered “meta-supported” if the p-value of association became smaller in the meta-analysis compared to that obtained in FGFP alone. To identify all independent SNPs associated with microbial traits, all meta-supported markers were clumped using plink and its default settings. The genome-wide meta-analysis p-value threshold was defined as 2.5×10^−08^ and the study-wide p-value threshold was defined as 1.57×10^−10^ using Bonferroni correction (assuming 2 million independent genetic association tests across 159 microbial traits).

Of the 153 microbial traits tested, two SNPs showed evidence of association that exceeded the study-wide meta-analysis p-value threshold and a further 11 exceeded the genome-wide meta-analysis threshold, where each SNP was associated with one microbial trait. These 13 SNPs were selected as genetic instruments for the 13 microbial traits within this two-sample MR analysis with overall and site-specific CRC, in addition to one SNP associated with bacteria of the *Bifidobacterium* genus that has previously been reported in the literature (**Supplementary Table 2**). Effect estimates from the mGWAS represent an increase in the log-transformed OR for P/A microbial traits and the standard deviation (SD) change for rank normalised AB microbial traits with each effect allele carried.

### Colorectal cancer GWAS data

Data for CRC were obtained from the most comprehensive GWAS of overall CRC to date from a meta-analysis of 120,328 individuals (comprising 55,168 CRC cases and 65,160 controls). The GWAS meta-analysis included the Genetics and Epidemiology of Colorectal Cancer Consortium (GECCO), Colorectal Cancer Transdisciplinary Study (CORECT), OncoArray + Custom, OmniExpress + Exome Chip, COloRectal cancer Study of Austria (CORSA) and UK Biobank (**Supplementary Table 1**) and full information on genotyping, imputation and QC have been described previously (32,33). Data for CRC sites were available across colon cancer (N=32,002), which is a sum of distal CRC, proximal CRC and cancer cases with unspecified sites, distal CRC (N=14,376), proximal CRC (N=15,706) and rectal cancer (N=16,212). CRC case status was diagnosed by a physician and approximately 92% of participants were white and of European ancestry. Data were obtained directly from the GECCO Coordinating Centre and ethics were approved by respective institutional review boards.

### Statistical analyses

#### Two-sample Mendelian randomization

In our primary analyses, we performed two-sample MR using the *TwoSampleMR*(34) package (version 0.5.6) in R to examine the causal relationship between 14 microbial traits and both overall and site-specific CRC risk.

Summary-level data (i.e., the SNP name [rsid], effect allele, other allele, effect allele frequency, beta coefficient, standard error, p-value and sample size) for each of the 14 microbial trait-associated SNPs associated were extracted from both the mGWAS and CRC GWAS meta-analysis. The proportion of variance explained (R^2^) in each microbial trait by each SNP and the strength of the instrument (assessed through the F-statistic) were calculated. The first was calculated using the following formula(35):

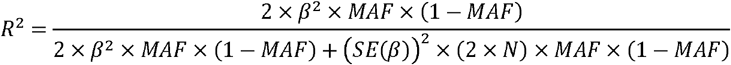

where *β* is the effect size (beta-coefficient), *MAF* is the minor allele frequency and *SE(*β) is the standard error of the effect size for a given SNP, and *N* is the sample size of the mGWAS. The second was calculated as follows:

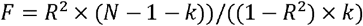

where *R*^*2*^ is the proportion of variance explained in the microbial trait by the SNP, *N* is the sample size of the GWAS, and *k* is the number of SNPs included in the instrument (i.e., for our main analysis, k=1 for each microbial trait)(36).

The exposure and outcome datasets were harmonized such that the effect of each SNP on the exposure and outcome was relative to the same effect allele. For ambiguously coded SNPs (i.e., “palindromic” SNPs where the effect/other allele were either an A/T or G/C combination), we used the effect allele frequency to resolve strand ambiguity, where possible. Non-inferable SNPs (i.e., “palindromic” SNPs with a MAF >0.42) were removed from the analysis.

Given that there was only one SNP associated with each microbial trait, the Wald ratio method was used as the main analyses, which estimates the effect of the exposure (here, each microbial traits) on the outcome (here, overall and site-specific CRC risk) by dividing the SNP-outcome association by the SNP-exposure association (**Figure 1**)(19).

### Sensitivity analyses

Three assumptions, namely (i) the relevance assumption (the genetic instruments used to proxy the exposure are strongly associated with that exposure), (ii) the independence assumption (there is no confounding between the genetic instrument(s) used to proxy the exposure and the outcome) and (iii) the exclusion restriction assumption (the genetic instruments used to proxy the exposure have no direct effect on the outcome other than through the exposure) must be met when assessing evidence for causal relationships between an exposure and outcome using MR(17–20). We explored possible invalidations of each of these assumption in our analyses by performing several sensitivity analyses that test the robustness of our findings and explored possible explanations of any observed causal effect via a process of elimination.

Firstly, genetic colocalisation was used to evaluate whether there was a shared causal variant at each locus responsible for variation in the gut microbiome and for conferring CRC risk, which is necessary but not sufficient for establishing causality. Secondly, in the absence of using formal statistical techniques that assess robustness of causal estimates owed to having only one SNP associated with each microbial trait, an alternative approach was used to test for any invalidation of the third MR assumption (i.e., the exclusion restriction assumption). We manually searched SNPs in two databases (PhenoScanner and the IEUOpenGWAS database) to identify traits they have previously been associated with. Thirdly, to formally test potential pleiotropy, we selected genetic variants to proxy microbial traits at a more lenient p-value threshold of 1×10^−05^ and repeated MR analyses with several complementary, pleiotropy-robust methods. Lastly, to assess whether any causal effect observed between microbial traits and CRC in our main analyses was indicative of reverse causation, we additionally performed two-sample MR in the reverse direction (i.e., with overall and site-specific CRC as the exposure and microbial traits as the outcome). Each of these sensitivity analyses are described in detail below.

### Colocalisation

Colocalisation analyses were conducted using the ‘coloc’ R package with default parameters(37). Bayes factor computation was used to generate 5 posterior probabilities (H0-H4) characterised by the following outcomes: (H0) neither trait has a genetic association in the region; (H1) only the gut microbiome has a genetic association in the region; (H2) only CRC has a genetic association in the region; (H3) both traits are associated but have different causal variants and (H4) both traits are associated and have the same causal variant. We used a posterior probability threshold ≥0.80 to indicate evidence of a shared common causal variant between each microbial trait and CRC. Full summary statistics were only available for the FGFP cohort for gut microbiome variation; therefore, genetic variants ±1Mb of the lead SNP associated with any microbial trait for which there was evidence for a causal impact on CRC in our main analyses were extracted from FGFP and GECCO genome-wide datasets. Regional association plots were generated to visualise genetic colocalisation using the LocusCompareR package(38).

### Manual exploration of pleiotropy

Where there was evidence for a causal effect of any microbial trait on CRC risk, summary-level GWAS data collated by PhenoScanner and the IEU Open GWAS were searched to identify whether any SNP used as an instrument for those microbial traits had been reported to be associated with CRC or any other trait that could be an independent cause of CRC. Given the number of results returned by PhenoScanner and the IEU Open GWAS at the time of searching (May 2022), a lenient p-value of 1×10^−04^ was set as the multiple testing threshold at which we defined evidence for an association between the SNP and any disease or trait(22,39)

### Two-sample MR using a lenient p-value threshold to select genetic instruments

To allow the use of more formal pleiotropy-robust methods that require multiple genetic instruments, a more lenient p-value threshold of 1×10^−05^ was used to expand the number of SNPs associated with each of the microbial traits in the main analyses(40,41). SNPs reaching this lenient p-value threshold were also then restricted to those which had directionally consistent effect estimates in each of the three studies in the mGWAS(21) (FGFP, FoCus and PopGen; see list of all directionally consistent SNPs in **Supplementary Table 14**). The IVW method was used and compared to the main analyses that utilised the Wald ratio estimator. The IVW method meta-analyses effect estimates across all SNPs weighted by the inverse variance of the SNP-outcome association using fixed effects. The caveats of using multiple SNPs defined at a more lenient p-value threshold in MR analyses are increasing the likelihood of weak instrument bias and introducing horizontal pleiotropic pathways between the SNPs and the outcome. Therefore, to test for horizontal pleiotropy in these sensitivity analyses, the weighted median(42), weighted mode(43) and MR-Egger(44) regression methods were also applied and consistency of effect estimates were compared to those obtained from the IVW method.

The weighted median(42) requires that only half the SNPs are valid instruments (i.e., exhibiting no horizontal pleiotropy, no association with confounders and a robust association with the exposure) for the causal effect estimate to be unbiased. The mode-based estimator clusters the SNPs into groups based on similarity of causal effects and returns the causal effect estimate based on the cluster that has the largest number of SNPs. The weighted mode(43) introduces an extra element similar to the IVW and weighted median estimators, weighting the contribution of each SNP to the clustering by the inverse variance of its outcome effect.

The MR-Egger(44) method is similar to the IVW approach but relaxes the “no horizontal pleiotropy” assumption. MR-Egger regression allows a non-zero intercept in the relationship between multiple SNP-outcome and SNP-exposure associations, where the intercept provides a formal statistical test for the presence of directional (bias inducing) horizontal pleiotropy. The slope of the MR-Egger regression between multiple SNP-outcome and SNP-exposure associations can be considered as an unbiased causal effect between the exposure (here, microbial traits) and the outcome (here, CRC), assuming any horizontal pleiotropic effects are not correlated with the SNP-exposure effects (i.e., strength of the instrument). Violations of the third MR assumption were also assessed by visual inspection of funnel(19), forest, scatter and leave-one-out plots, and tests of heterogeneity(18) of effects between the SNPs using Cochan’s Q statistic(45).

Effect estimates obtained from all two-sample MR analyses represent the OR for both overall and site-specific CRC risk for each SD higher abundance of each continuous microbial trait (those denoted with “AB” in tables) and the OR for CRC for an approximate doubling of the genetic liability to presence (vs. absence) of each binary microbial trait (those denoted with “P/A” in tables)(46). For all analyses, P-values were interpreted as continuous indicators of evidence strength and conclusions were drawn based on effect sizes and their precision. Given the high correlation between microbial traits, there was no correction for multiple testing. Analyses and data visualisation were conducted using the *TwoSampleMR* R package (version 0.5.5) and the ieugwasr R package (0.1.5) in with R (version 4.1.0) and PhenoScanner (version 2) online(22,34,39).

### Reverse two-sample Mendelian randomization

For reverse MR analyses, 57 SNPs that reached the conventional genome-wide p-value threshold (P<5×10^−08^) were selected as instruments for overall CRC risk, obtained from a large GWAS meta-analysis of CRC comprising 34,627 CRC cases and 71,379 controls of European ancestry(47). For CRC sites, 109 SNPs associated with each of the four CRC sites (P<5×10^−08^) were obtained from the largest and most recent GWAS meta-analysis of anatomical site-specific loci(32). Summary statistics for CRC-associated SNPs were extracted for the 14 microbial traits from the FGFP cohort (n=2223) from the Hughes *et al*., (2020) GWAS(21). MR analyses were undertaken in the same was as described above, with the IVW method used as the main analyses, results of which were compared to those obtained from the MR-Egger, weighted median, and weighted mode-based estimators that test the assumptions of no pleiotropy among genetic instruments and outcomes.

## Data Availability

GWAS summary-level data used in this study were publicly available for microbiome GWAS (mGWAS) conducted by Hughes et al. 2020. Summary-level data for overall and site-specific CRC were obtained directly from GECCO (summary-level data are publicly available for the CRC GWASs used in the reverse MR analyses.

## Author contributions

KHW conceived the study with LHN, ADJ, RCT, ATC and MG. KHW, CH and GR performed the data analysis. YL and CQ provided access to GECCO data. DAH and JR provided access to FGFP data. KHW, CH and GR were involved in original manuscript preparation. All authors critically reviewed the manuscript and contributed important intellectual content. All authors have read and approved the final manuscript as submitted.

## Data availability

GWAS summary-level data used in this study were publicly available for microbiome GWAS (mGWAS) conducted by Hughes *et al*. 2020(21). Summary-level data for overall and site-specific CRC were obtained directly from GECCO (summary-level data are publicly available for the CRC GWASs used in the reverse MR analyses (47) (32).

